# Preliminary clinical characteristics of Pediatric Covid-19 cases during the ongoing Omicron XBB.1.16 driven surge in a north Indian city

**DOI:** 10.1101/2023.04.18.23288715

**Authors:** Vipin M. Vashishtha, Puneet Kumar

## Abstract

India is experiencing a new surge in Covid-19 cases in most parts of the country. A new sub-variant of Omicron, XBB.1.16 which is far more aggressive and immune evasive than other sub-lineages of Omicron, is responsible for this outbreak. In this preliminary account, we describe key clinical characteristics of SARS-CoV-2 infected children, visiting an outdoor department of a pediatric hospital in a north Indian city. Our preliminary findings show a higher involvement of young infants than older children and mild respiratory illness predominates other presentations. One interesting finding was the presence of itchy, non-purulent conjunctivitis with mucoid discharge and stickiness of eyelids in 42.8% of positive infants. None of the children required hospitalization. All recovered with symptomatic treatment.

**Key findings:**

- The current ongoing XBB.1.16 driven surge of Covid-19 is causing mild febrile illness in children in India
- Young infants are disproportionately more affected than older children.
- Unlike the previous BA.2 Omicron wave, respiratory symptoms are predominating the clinical presentation in young infants in the ongoing surge.
- Conjunctival involvement is seen in 42.8% of affected infants.

## Introduction

A new surge of Covid-19 cases is seen in India since the first week of March 2023. The number of daily cases as well as active cases have been steadily rising with more than 11,000 cases reported on April 14, 2023, and the active case tally rose to around 50,000 **(1)**. Cases are also steadily increasing in Uttar Pradesh, the most populous state of India since early March. A new sub-lineage of the Omicron variant of Severe Acute Respiratory Syndrome Corona Virus-2 (SARS-CoV2) called XBB.1.16 is behind this surge **(2)**. According to the most recent INSACOG data, the percentage of this variant is currently 58.6 % of all the isolates sequenced and uploaded to this site **(3)**.

### Evolution of Omicron VOC and the emergence of XBB.1.16 in India

Starting from late November 2021 when the Omicron (B.1.1.529) was first detected, it led to big surges globally in January 2022, and eventually became the most dominant variant globally, eking out the previous dominant variant Delta (B.1.617.2). After large wave of Omicron BA.2 in January, smaller surges were caused by BA.2 sub-lineages, mainly BA.2.38 and BA.2.75 from April to June 2022 **(3)**. Other key sub-lineages of Omicron like BQs, BA.4, BA.5, and BF.7 could not generate any significant surge in cases in a large part of the country despite leading to large waves of infections in many countries around the world. The XBB sub-lineage, a recombinant of the BA.2.10.1 and BA.2.75 sub-lineages and suspected to be one of the most immune-evasive strains, was first identified in India in August 2022 and has since spread rapidly around the world **(4)**. From January 2023, the XBB and its sub-lineages gradually replaced the other dominant sub-lineage BA.2.75 completely.

**Figure 1.**
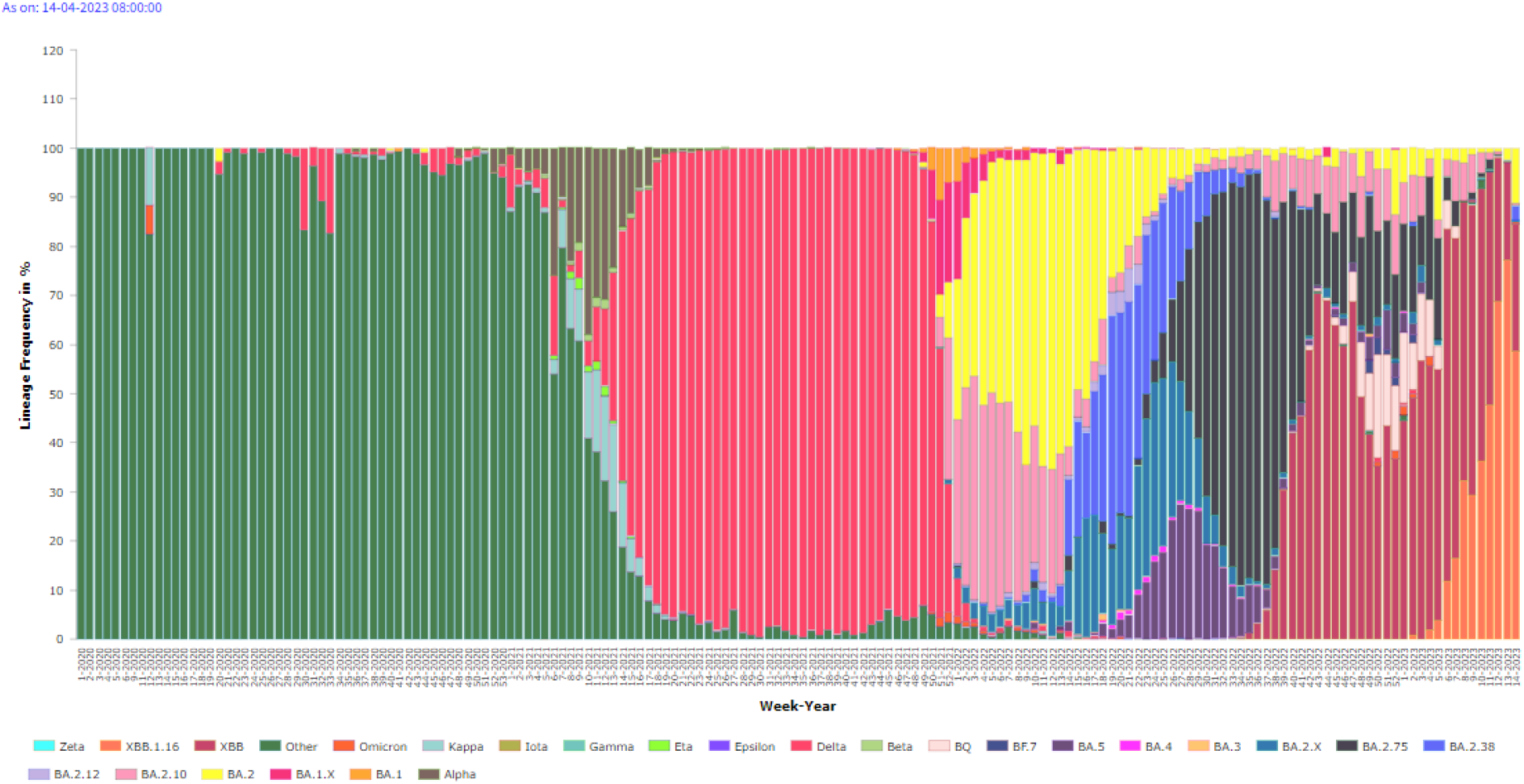
Various lineages of Severe Acute Respiratory Syndrome Corona Virus-2 (SARS-CoV2) at different time point in India (Source: INSACOG)

In late February, XBB.1.16 (“Arcturus”) an XBB.1 offspring emerged in India and outcompeted all other variants by end of March 2023. A big surge is being currently observed, with test positivity touching 6.78% and crossing even 30% in some states like Delhi. With its rapid spread to more than 25 countries, the World Health Organization (WHO) on March 22, 2023, dubbed it as the ‘variant under monitoring’ **(5)**.

Interestingly, the other XBB descendent, XBB.1.5 became the globally dominant sub-variant and under the WHO’s tag of ‘variant of interest’, could not find its foothold in India **(5)**. Compared to XBB.1.5, XBB.1.16 has two substitutions in the Spike protein: E180V in the 55 N-terminal domain, and T478R in the receptor-binding domain (RBD) **(6)**. Many experts believe that XBB.1.16 has the potential to become a global dominant variant replacing all other circulating sub-lineages **(7)**.

Notably, XBB.1.16 has an effective reproductive number (Re) that was 1.27- and 1.17-fold higher than the parental XBB.1 and XBB.1.5, respectively. Virological studies have also shown the binding affinity of XBB.1.16 RBD to human ACE2 receptor is higher than that of XBB.1. As with previous sub-variants of Omicron, it also has a combination of mutations in spike protein: one that increases infectivity (S: T478R substitution) and another that evades antiviral immunity but attenuates infectivity (S: E180V substitution). Neutralization assays demonstrated the robust resistance of XBB.1.16 to breakthrough infection sera of BA.2 (18-fold versus B.1.1) and BA.5 (37-fold versus B.1.1). Yamasoba et al **(8)** tested six clinically available monoclonal antibodies and showed that XBB subvariants, including XBB.1.16, are resistant against all except sotrovimab. They suggested that the greater growth advantage of XBB.1.16 as compared to XBB.1 and XBB.1.5, along with more profound immune evasion may be due to (1) different antigenicity from XBB.1.5; and/or (2) the mutations in the non-S viral protein(s) that may contribute to increased viral growth efficiency **(8)**.

## Methods

This single-centre, observational descriptive study is being conducted at a secondary level, exclusive pediatric hospital of western Uttar Pradesh, India since April 4, 2023. An early account of clinical characteristics of pediatric Covid-19 cases presented till April 16, 2023, is being presented in this report.

All outdoor pediatric cases visiting this hospital with a mild respiratory, or febrile illness were asked to undertake a SARS-CoV-2 RT-PCR or RAT. A copy of the prescription of the child visiting the outpatient department (OPD) and undergoing these tests and suspected to be SARS-CoV-2 infected was preserved. We recorded the key demographic characteristics like age, sex, weight, contact history, presenting signs and symptoms, and comorbidities of all these children. Positive Covid-19 cases are grouped into three cohorts based on their age: infants (0-11 months), young children (12-59 months) and older children (above 60 months).

A reading of above 99.5 °F was considered evidence of fever according to Facility Based Integrated Management of Neonatal and Childhood Illness (F-IMNCI) **(9)**. Both RAT and RT-PCR were employed to confirm a case. However, all positive samples were reanalysed with RT-PCR to assess cycle threshold (Ct) values. Ethical approval to perform this study has been approved by the Institutional Ethics Committee, Mangla Hospital & Research Centre, Bijnor, Uttar Pradesh, India, which is affiliated with the Institutional Review Board of Christian Medical College, Vellore, India (Ethics approval letter: MHRC/IEC/2023/03 dated April 3, 2023).

## Results

Out of 127 children with suspected Covid-19 features who presented at the OPD, the parents of 90 could get the RAT or RT-PCR for the SARS-CoV-2 test. Twenty-five cases came positive for SARS-CoV-2 infection **(Table I**). Of these 25 positive cases, none required hospitalization for their illness.

**Table I.**
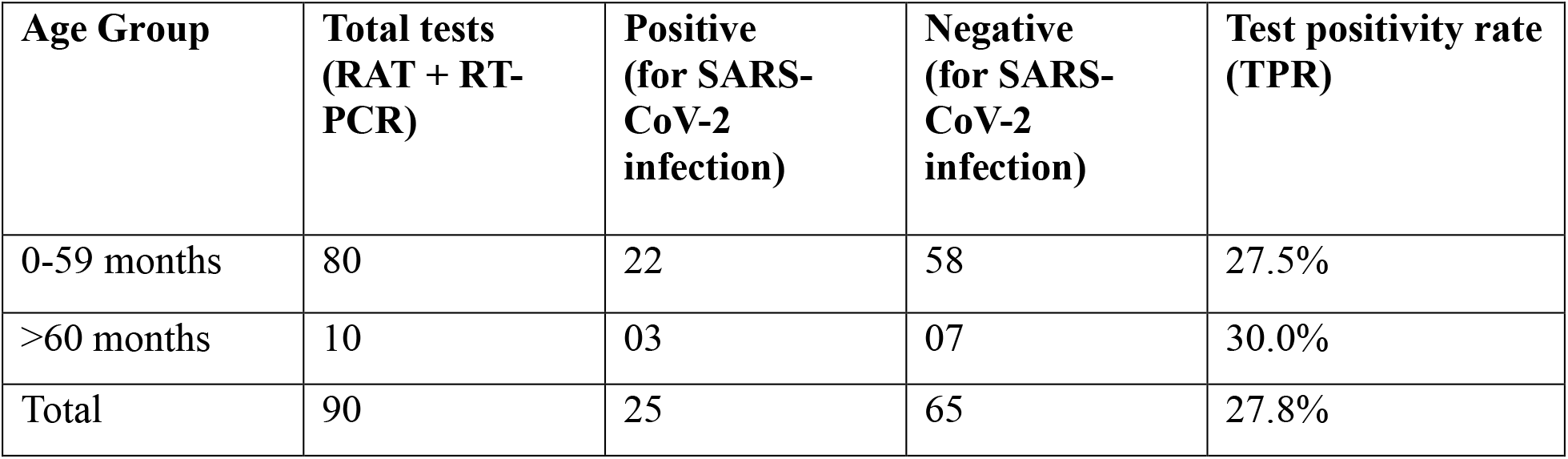
SARS-CoV-2 infection-positive cases and test positivity rate in different age groups.

In the SARS-CoV-2 positive cases, the total duration of acute illness was 1-3 days. On sub-group analysis, it was noted that infants below one year had a significantly higher positivity rate than older children (40.38% versus 10.5%, p=000816). Mild to moderate fever (100-102°F) was noticed in all but two infants. Acute respiratory symptoms predominated the Covid-19 illness in all except six children in all three cohorts. Expectedly, symptoms like headache, muscle pains and body aches, throat pain were seen exclusively in older children since it was not possible to obtain the history of these subjective symptoms from young children. None of the older children complained of anosmia or loss of taste **(Table II)**. The cycle threshold values for nucleocapsid and ORF1ab genes on positive RT-PCR vary from 18 to 23.

**Table II.**
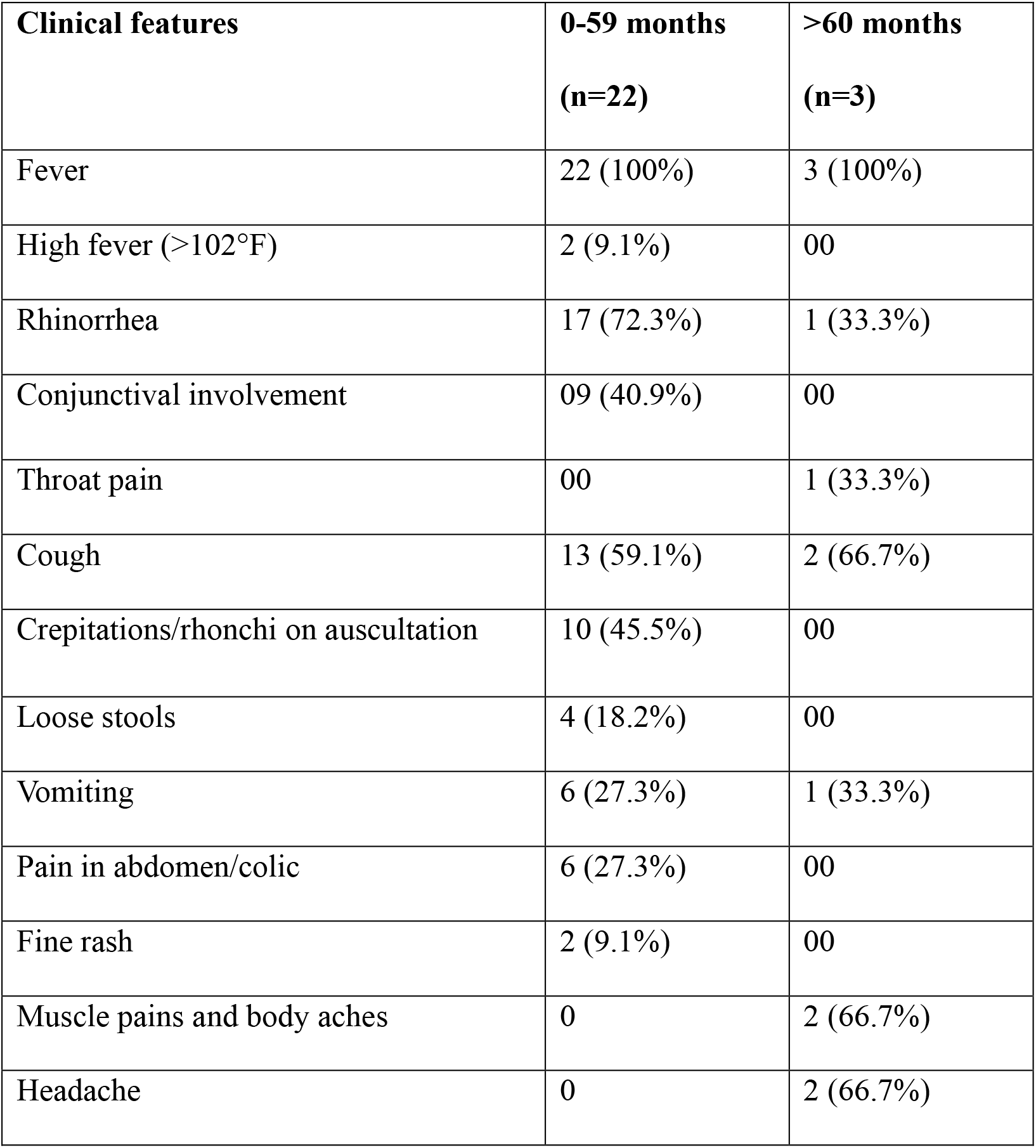
Clinical Characteristics of young and older children with COVID-19 Illness.

Although, we could notice a fine, erythematous rash in two infants, no case resembling the features of MIS-C was seen. Surprisingly, only the parents of three positive cases could give a history of acute illness in any of the family members.

One interesting finding of the study was the presence of itchy, non-purulent conjunctivitis with mucoid discharge and sticky eyelids in nine infants (42.85%). This manifestation was not seen amongst infants during the past Covid-19 waves, especially in this region. None of the children required hospitalization. All recovered with symptomatic home-based treatment.

## Discussion

In this brief review, a preliminary account of the clinical features of pediatric Covid-19 cases appearing during the ongoing Covid-19 surge is presented. Though we could not perform genetic sequencing, we believe that most positive cases represent XBB.1.16 sub-variant infection. Like the third wave of Covid-19 in India, the current ongoing outbreak is also causing mild brief symptomatic illness in children **(10, 11)**. Most affected children only have a mild respiratory illness characterized by fever, cold, cough and bronchitis that resolves within one to three days. However, the disease is found to be more prevalent in infants than in older children. Unlike the previous BA.2 Omicron wave in India in which the gastrointestinal symptoms predominated in young infants, mainly respiratory symptoms are dominating the clinical picture of the ongoing surge so far **(10)**.

One surprising finding, not seen in the previous waves, is the presence of itchy, non-purulent, conjunctivitis that was solely seen in infants. Although, frank conjunctivitis was a common clinical presentation of Covid-19 during the early part of its emergence in some regions **(12)**. However, later, this clinical sign was not considered to be a key finding of Covid-19. Many respiratory viruses like adenovirus, influenza, RSV, measles, etc are more commonly associated with conjunctival involvement **(13)**. However, unlike in these viral infections, the conjunctival affliction was mild and not associated with marked redness and purulent discharge in the current outbreak. Furthermore, we did an adenovirus RT-PCR in two such infants which came negative.

One limitation of the current study is the proper case selection since we only tested the symptomatic children. We find strong reluctance among parents to let their children be tested for SARS-CoV-2.

## Conclusions

Though the current ongoing Omicron’s XBB.1.16-driven surge of Covid-19 is causing mild illness in children, the SARS-CoV-2 virus continues to evolve at a rapid pace. The scientific community needs to remain proactive in monitoring this evolution, especially regarding the transmissibility, immune evasiveness, pathogenicity, and resistance to existing anti-viral medications to provide guidance to all stakeholders to launch appropriate strategies to counter the growing threat. The study findings may benefit clinicians, paediatricians, public health experts and parents for early identification of the disease and proper management.

## Data Availability

All data produced in the present work are contained in the manuscript

## Contributors

VMV conceptualize and conducted the study, analysed, and prepared the initial draft of the manuscript. PK helped in drafting, and data analysis and gave valuable inputs in the interpretation of the data and helped in editing the manuscript. Both authors approved the final version of the manuscript.

## Acknowledgements

*We are grateful for the cooperation of all participants, parents/guardians, and laboratory staff of our centre*.

## REFERENCES

1- uge jump in COVID-19 cases! Over 11,000 infections reported in last 24 hours. Available online: https://www.businesstoday.in/coronavirus/story/huge-jump-in-covid-19-cases-over-11000-infections-reported-in-last-24-hours-377357-2023-04-14 [Last accessed on 2023 Apr 15]

2- XBB.1.16, new Arcturus variant behind India’s COVID-19 surge. Available online: https://health.economictimes.indiatimes.com/news/industry/xbb-1-16-new-arcturus-variant-behind-indias-covid-19-surge/99495285 [Last accessed on 2023 Apr 15]

3- The Indian SARS-CoV-2 Genomics Consortium (INSACOG). Available online: https://inda.rcb.ac.in/insacog/statreportlineagegraph [Last accessed on 2023 Apr 16]

4- Goh AXC, Chae SR, Chiew CJ, Tang N, Pang D, Lin C, et al. Characteristics of the omicron XBB subvariant wave in Singapore. Lancet. 2023 Apr 15;401(10384):1261–1262.

5- Lee BY. XBB.1.16 ‘Arcturus’ Is New Covid-19 Variant Under Monitoring By The WHO. Available online: https://www.forbes.com/sites/brucelee/2023/04/16/xbb116-arcturus-is-new-covid-19-variant-under-monitoring-by-the-who/?sh=2b7882c04224 [Last accessed on 2023 Apr 17]

6- Mathur N. The virological characteristics of XBB.1.16. Available online: https://www.news-medical.net/news/20230412/The-virological-characteristics-of-XBB116.aspx [Last accessed on 2023 Apr 16]

7- Uniyal P. All about XBB.1.16, new Omicron variant behind India’s Covid-19 spike, from symptoms to risk factors. Available online: https://www.hindustantimes.com/lifestyle/health/all-about-xbb-1-16-new-omicron-variant-behind-india-s-covid-19-spike-from-symptoms-to-risk-factors-101680160372702.html [Last accessed on 2023 Apr 16]

8- Yamasoba D, Uriu K, Plianchaisuk A, et al. Virological characteristics of the SARS-CoV-2 Omicron XBB.1.16 variant. BioRxiv. Available online: https://www.biorxiv.org/content/10.1101/2023.04.06.535883v3.full [Last accessed on 2023 Apr 16]

9- Molteni E, Sudre CH, Canas LS, et al. Illness duration and symptom profile in symptomatic UK school-aged children tested for SARS-COV-2. Lancet Child Adolesc Health. 2021;10:708–18.13.

10- Sarkar M, Ghosh A, Konar MC, Roy O, Mahapatra MK, Nandi M. Clinical Characteristics of Children With SARS-CoV-2 Infection During the Third Wave of the Pandemic: Single Center Experience. Indian Pediatr. 2022 Jul 15;59(7):531–534.

11- Muthusamy S, Sarojam B, Sugunan S, Krishna G S B A S AK. Clinical Profile and Short-Term Outcome of Children with Acute SARS-CoV-2 Infection During the First and Second Waves of the Pandemic. Indian J Pediatr. 2022 Jun 23:1–7.

12- Güemes-Villahoz N, Burgos-Blasco B, García-Feijoó J, Sáenz-Francés F, Arriola-Villalobos P, Martinez-de-la-Casa JM, et al. Conjunctivitis in COVID-19 patients: frequency and clinical presentation. Graefes Arch Clin Exp Ophthalmol. 2020 Nov;258(11):2501–2507.

13- Belser JA, Rota PA, Tumpey TM. Ocular tropism of respiratory viruses. Microbiol Mol Biol Rev. 2013 Mar;77(1):144–56.

